# Neuroanatomic markers of post-traumatic epilepsy based on magnetic resonance imaging and machine learning

**DOI:** 10.1101/2020.07.22.20160218

**Authors:** Haleh Akrami, Richard M. Leahy, Andrei Irimia, Paul E. Kim, Christianne N. Heck, Anand A. Joshi

## Abstract

Although post-traumatic epilepsy (PTE) is a common complication of traumatic brain injury (TBI), the relationship between these conditions is unclear, early PTE detection and prevention being major unmet clinical challenges. This study aims to identify imaging biomarkers that distinguish PTE and non-PTE subjects among TBI survivors based on a magnetic resonance imaging (MRI) dataset. We performed tensor-based morphometry to analyze brain shape changes associated with TBI and to derive imaging features for statistical group comparison. Additionally, machine learning was used to identify structural anomalies associated with brain lesions. Automatically generated brain lesion maps were used to identify brain regions where lesion load may indicate an increased incidence of PTE. Statistical analysis suggests that lesions in the temporal lobes, cerebellum, and the right occipital lobe are associated with an increased PTE incidence.

## 1. Introduction

The onset of post-traumatic epilepsy (PTE) after traumatic brain injury (TBI) is relatively common [40]. Epidemiological studies have found that PTE accounts for 10-20% percent of all symptomatic epilepsies in the general population and ∼5% of all epilepsies [20, 43]. Significant risk factors for seizure onset over one week after TBI include the occurrence of seizures within the first week, acute intra-cerebral (especially subdural) hematoma, brain contusion, greater injury severity, and age over 65 at the time of injury [9]. As many as 86 percents of patients with one seizure early after TBI experience a second one within the next two years [33].

Despite the reported relationship between TBI and PTE, identifying biomarkers of epileptogenesis after TBI is still a fundamental challenge. Preliminary studies in adult male Sprague-Dawley rats indicate the potential involvement of the perilesional cortex, hippocampus, and thalamus in PTE and demonstrated the potential of leveraging MRI analysis to find PTE biomarkers [23, 44]. Previous MRI studies have shown correlations between PTE incidence and (a) the presence of lesions in *T*_2_-weighted scans, (b) injury severity and (c) injury type [4, 12, 15, 42]. Studies of PTE reported correlations between PTE and the existence of frontal, parietal, and temporal lesions [47, 1, 48, 16, 50]. Nevertheless, the association between PTE and lesion size or location remains poorly understood. Additionally, the heterogeneous nature of TBI injury types, pathology, and lesions present additional challenges to biomarker discovery. Because the locations, spatial extent, and content of lesions vary considerably between patients with vs. without PTE, there is no complete spatial overlap of injury profiles across the two groups. This heterogeneity needs to be accounted for in statistical analyses due to its potentially confounding effect. The prediction of post-traumatic seizure onset and frequency based on neurological and radiological examinations has been only moderately successful and more research is needed to understand the relationship between TBI and PTE [46, 45, 17, 14, 24]. Thus, the identification of imaging biomarkers can help in developing better PTE prediction strategies.

This study uses multimodal MRI to identify location- and contrast-related biomarkers of PTE. We perform two analyses aimed at characterizing changes in brain structure using two distinct strategies:

- *Morphometric Analysis:* We performed a population analysis of morphometric changes in the brain associated with TBI. In contrast to the lesion analysis described below, this analysis focuses on identifying changes in brain shape rather than alterations in tissue composition.
- *Lesion analysis:* We use a machine learning (ML) method for identifying abnormal contrasts in multimodal MR images, which are indicative of lesions and tissue abnormalities such as edema, hematoma, and hemorrhage.

## 2. Materials and methods

### 2.1. Data

We used three datasets in this study: 1) the Maryland TBI MagNeTs dataset [19], including 74 subjects used for statistical comparison and the remaining 41 subjects (total 114) were used for training the neural network;2) the TRACK-TBI Pilot [52] dataset including 97 subjects were used for training the neural network; and 3) the ISLES (The Ischemic Stroke Lesion Segmentation) dataset with manually delineated lesions [39], were used as validation data for measuring the accuracy of our lesion delineation method. Information about these three datasets is provided below.

#### 2.1.1. Maryland MagNeTs data

The total number of subjects available from this dataset was 115. The dataset was collected as a part of a prospective study that includes longitudinal imaging and behavioral data from TBI patients with a Glasgow Coma Scores (GCS) in the range of 3-15 (mild to severe TBI). Injury mechanisms included falls, bicycle or sports accidents, motor vehicle collisions, and assaults. The individual or group-wise GCS, injury mechanisms, and clinical information is not shared. The imaging data are available from FIT-BIR (https://fitbir.nih.gov), with FLAIR, *T*_1_, *T*_2_, diffusion, and other modalities available for download. In this study, we used imaging data acquired within 10 days after injury, and seizure information was recorded using follow-up appointment questionnaires. Exclusion criteria included a history of white matter disease or neurodegenerative disorders including multiple sclerosis, Huntington’s disease, Alzheimer’s disease, Pick’s disease, and a history of stroke or brain tumors.

Imaging was performed on a 3T Siemens TIM Trio scanner (Siemens Medical Solutions, Erlangen, Germany) using a 12-channel receiver-only head coil. For statistical analysis, we used 37 subjects with epilepsy (26M/11F) from this dataset and 37 randomly selected subjects without epilepsy (distinct from the set used to train the lesion detection algorithm; 27M/10F) from the same dataset [19]. The age range for the epilepsy group was 19-65 years (yrs) and 18-70 yrs for the non-epilepsy group. The analysis of population differences was performed using the *T*_1_-weighted, *T*_2_-weighted and FLAIR MRIs in the Maryland TBI MagNeTs dataset [19]. The remaining 41 subjects with TBI but without PTE used for training the machine learning algorithm.

#### 2.1.2. TRACK-TBI Pilot dataset

This is a multi-site study with data across the injury spectrum, along with CT/MRI imaging, blood biospecimens, and detailed clinical outcomes [52]. Patients were scanned at 3T and their information was collected according to the 26 core Common Data Elements (CDEs) standard developed by the neuroimaging working group, including 93 CDEs. The 3T research MRI scanners were manufactured by General Electric, Phillips, and Siemens. The MRI protocols complemented the CDE tiers used in the Alzheimer’s Disease Neuroimaging Initiative (ADNI) with *T*_*R*_*/T*_*E*_ = 2200*/*2.96 ms, an effective *T*_*I*_ of 880 ms, an echo spacing time of 7.1 ms, a bandwidth 240*Hz/pixel*, a total scan time of 4 minutes and 56 seconds. The *T*_1_-weighted sequence adapted for non-ADNI studies was adopted because the MP-RAGE (magnetization-prepared rapid gradient-echo) sequence with 180-degree radio-frequency pulses was a key component of the ADNI MRI protocol and increased the probability of obtaining at least one high-quality morphometric scan at each examination.

To train the machine learning algorithm, we used 2D slices of brain MRIs from a combined group of 41 subjects (33M/8F, age range 18-82 yrs) with TBI but without PTE from the Maryland TBI MagNeTs study (excluding the 74 subjects used for statistical testing) [19] and 97 subjects with TBI but without epilepsy from the TRACK-TBI Pilot study [52] (70M/27F, age range 11-73 yrs), available for download from https://fitbir.nih.gov.

#### 2.1.3. ISLES dataset

For validation of the ability of the machine learning algorithm to identify lesions, we used 15 subjects from the ISLES (The Ischemic Stroke Lesion Segmentation 2015) database. The data is available for download at http://www.isles-challenge.org/ISLES2015. This dataset was used because it provides manually delineated lesions as ground truth [39]. The data consists of T1, T2, and FLAIR images as well as manual delineations of the lesions. Imaging was performed on 3T scanners manufactured by Phillips systems with routine clinical acquisition parameters.

### 2.2. Pre-processing

The pre-processing of all three datasets was performed using the Brain-Suite software (https://brainsuite.org). The three modalities (*T*_1_, *T*_2_, FLAIR) were co-registered to each other by registering *T*_2_ and FLAIR to *T*_1_, and the result was co-registered to the MNI atlas by registering *T*_1_ images to the MNI atlas using a rigid (translation, scaling and rotation) transformation model. As a result, we have all three modality images in MNI space at 1 mm^3^ resolution. Skull and other non-brain tissue were removed using BrainSuite. Brain extraction was performed by stripping away the skull, scalp, and any non-brain tissue from the image. This was followed by tissue classification and generation of the inner and pial cortex surfaces. Subsequently, for training and validation, all images were reshaped into 128 × 128 pixel images and histogram-equalized to a lesion free subject.

The extracted cortical surface representations and brain image volumes for each subject were jointly registered to the BCI-DNI Brain Atlas (http://brainsuite.org/svreg_atlas_description/) using BrainSuite’s Surface-Volume Registration (SVReg18a) module [29, 28]. SVReg uses anatomical information from both the surface and volume of the brain for accurate automated co-registration, which allows consistent surface and volume mapping to a labeled atlas. This co-registration establishes a one-to-one correspondence between individual subjects’ *T*_1_ MRIs and the BCI-DNI brain atlas. The deformation map between the subject and the atlas encodes the deformation field that transforms images between subject and atlas.

### 2.3. Tensor-based morphometry

To perform a morphometric analysis that compares the brain shapes of PTE patients to those of non-PTE participants, we used tensor-based morphometry (TBM) [11, 22]. TBM is an established neuroimaging method that identifies regional differences in brain structure in groups or individuals relative to a control group using the determinant of the Jacobian matrix computed from the deformation field; the latter defines a nonlinear mapping that warps the brain into a common (atlas) space [5, 38]. Regions of the brain that differ most from the reference atlas brain will be characterized by significantly smaller (e.g. atrophy/tissue loss) or larger (e.g. enlarged ventricles) Jacobian determinants relative to healthy controls (HCs).

We used Brainsuite’s TBM pipeline to map structural brain changes resulting from TBI to identify regions that are more strongly associated with the onset of PTE [21, 11]. The Jacobians are computed from the deformation fields associated with the cortically constrained volumetric subject-to-atlas registration described above. We applied 3-mm standard deviation (7 mm full width at half-maximum, or FWHM) isotropic smoothing to the Jacobian determinant maps to account for residual misregistration and to increase statistical power.

We analyzed the Jacobian determinants at each voxel in two ways: (1) a *t*-test to determine if there are differences in the mean local shape across the PTE and non-PTE groups, (2) an F-test to determine if there are group differences in the variances of this measure. The null hypothesis for (1) was that the mean of the Jacobian determinants is the same in the PTE and non-PTE groups. For (2), the null hypothesis was that the variance of Jacobian determinants in the PTE and non-PTE groups is the same.

The *t*-test would reveal if there are consistent TBI-related brain shape differences between the PTE and non-PTE groups. Since trauma affects different areas of the brain in different subjects across groups, it seems unlikely that consistent localized differences between the two groups should be observed. For this reason, we also included the F-test, which would allow us to observe larger variances in localized shape differences in the PTE group in regions at higher risk for developing PTE foci. Since there may be more than one such region, only a subset of PTE subjects would have TBI-related variance difference in any particular area, leading to a larger variance across the PTE group in these areas relative to the non-PTE group.

### 2.4. Lesion-based analysis

To complement the TBM analysis*-*which captures morphometric brain changes*-*we also performed a lesion-based analysis to analyze changes in the underlying tissue microstructure, edema, and other TBI-related factors revealed by MRI contrast changes. For lesion mapping, we use multimodal MR images (*T*_1_, *T*_2_, FLAIR) and machine learning (ML) to automatically identify and delineate abnormal tissues. Lesions can be identified by visual inspection after extensive training, but this time-consuming process makes ML an attractive alternative. Approaches based on supervised ML have already achieved noticeable success, reaching high accuracy for lesion detection [37, 30, 41]. However, many manual lesion delineations are required to train supervised machines. Unsupervised approaches, on the other hand, do not require labeled data but are typically less accurate.

A popular unsupervised ML approach to lesion identification leverages a form of deep learning (DL) neural network known as a variational autoencoder (VAE) [7, 31]. By training the VAE using nominally normal imaging data, the network learns to encode only normal images. As a result, the associated image ‘decoder’ can reconstruct normal images. When presented with images containing lesions or anomalies, the VAE encodes and reconstructs the image as if it contained only normal structures, as illustrated in Figure 1. Lesions can then be identified from the differences between original and VAE-decoded images. In practice, VAEs exhibit some degree of robustness to outliers (in our case, lesions) in the training data.

**Figure 1:**
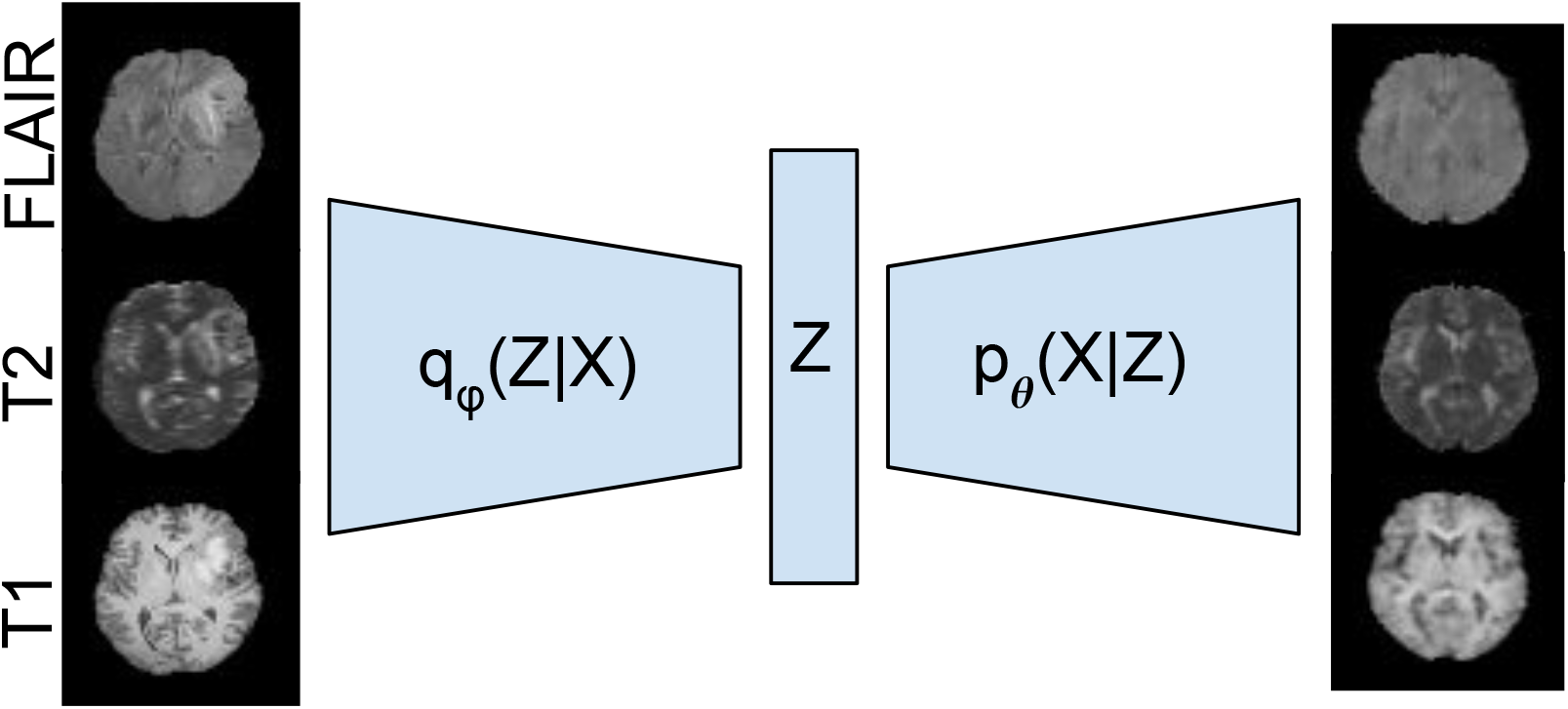
The VAE network and an input/output sample pair from the ISLES dataset. 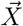 denotes the input data, 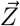 denotes its low-dimensional latent representation. The VAE consists of an encoder network that computes an approximate posterior 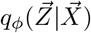, and a decoder network that computes 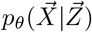. The VAE model takes *T*_1_, *T*_2_, and FLAIR images from individual subjects (left), compresses them to generate a latent representation (*Z*), and regenerates three images (right). The VAE is trained on a dataset that contains few lesions. After training, when presented with a new lesioned brain, the reconstruction effectively removes the lesion from the image resulting in a normal (lesion-free) version of the brain.

To identify lesion-based PTE biomarkers, we trained the VAE using the *T*_1_-weighted, *T*_2_-weighted, and FLAIR MRIs in the Maryland TBI MagNeTs dataset [19]. The lesions were delineated based on VAE reconstruction error in the FLAIR images. We used the VAE for unsupervised delineated of lesions in these two groups. A detailed description and validation of our method are available elsewhere [2]. The VAE is a directed probabilistic graphical model whose posteriors are approximated by a neural network.

Since a VAE learns a general pattern in the training data, its encoder does not encode the lesion and its decoder reconstructs a lesion-free version of the image. The difference between the input and output (reconstructed) images can then reveal the locations of lesions as well as other pathology which may include hematoma, edema, and hemorrhage. We used a combination of the Maryland TBI MagNeTs data and TRACK-TBI Pilot data to train our VAE. These datasets are not lesion-free but a VAE can handle occasional lesions in the training set since it has some robustness. To ensure this, we compared its performance to our proposed robust VAE [2, 3] and confirmed that there was no significant difference between their results. In the following, we refer to this procedure of computing differences of input and reconstructed brains as ‘lesion mapping’. We then investigate the relationship between the sizes and locations of lesions and PTE onset.

For the VAE’s architecture, we used the convolutional neural network (CNN) proposed in [32] that consists of three consecutive blocks of convolutional layers, a batch normalization layer, a rectified linear unit (ReLU) activation function and two fully-connected layers in the bottleneck for the encoder. For the decoder, we used a fully-connected layer and three consecutive blocks of deconvolutional layers, a batch normalization layer and a ReLU with a final deconvolutional layer (Figure 1). The size of the input layer is 3 × 128 × 128. The VAE error indicates deviation from normal tissue and therefore we refer to it as VAE lesion map, was then warped to the BCI-DNI atlas space by applying the deformation field computed during the registration process to the atlas during preprocessing.

Similar to the TBM analysis, we analyzed the VAE lesion maps using two methods: (1) a *t*-test of whether there are differences in the means of VAE lesion maps between the PTE and non-PTE groups, (2) an F-test to determine whether there are statistically significant differences in the variances of lesion maps between the PTE and non-PTE TBI groups.

Additionally, we performed a regional analysis by quantifying lesion volume from binarized lesion maps in each lobe using our USCLobes brain atlas (http://brainsuite.org/usclobes-description). This atlas consists of lobar delineations (left and right frontal, parietal, temporal, and occipital lobes, as well as the bilateral insulae and cerebellum). To identify the lesions as binary masks in each lobe, a one-class support vector machine (SVM) [53] applied to the VAE lesion maps at each voxel and across subjects to identify subjects with abnormally large errors at that voxel. One-class SVM is a commonly used unsupervised learning algorithm for outlier detection [53, 13]. We used the outliers marked by the one-class SVM as lesion delineations and computed lesion volumes per lobe by counting the number of outlier voxels in each lobe for each subject.

Since lesion locations vary across subjects, some subjects in either group have healthy-appearing tissue at a given location in the brain whereas some have lesions. However, if lesions in a brain region increase the chance of PTE, then in that region, we would expect to see a greater heterogeneity of image intensities across the PTE than in the non-PTE group, leading to an increase in variance. To test this hypothesis at each voxel, we performed an F-test on the lesion maps. The resulting p-values were corrected for multiple comparisons using Benjamini and Hochberg’s false discovery rate (FDR) procedure [6].

#### 2.4.1. Validation of VAE lesion detection

After training the VAE, we evaluated its performance using the ISLES dataset. We calculated the pixel-wise absolute reconstruction error and applied median filtering to the resulting image to remove isolated pixels. Ground truth was defined using hand-traced delineation of lesions on FLAIR images [39]. We then generated ROC (receiver operating characteristics) curves and computed the AUC (area under the curve) based on concordance between pixels in the labeled lesions and those pixels in which the absolute error image exceeded a given threshold. The ROC curves were generated by varying the lesion threshold intensity in the error image to control the true and false-positive rates. We also generated reconstruction error maps for the Maryland MagNeTs test set and used these for the lesion mapping analysis described above.

## 3. Results

### 3.1. TBM-based analysis

The results of TBM analysis using a *t*-test of Jacobian determinant maps are shown in Figure 2 while the results for the F-test are shown in Figure 3. As anticipated, in the case of the *t* statistic map, TBM analysis results did not survive multiple comparison corrections for the false discovery rate (FDR) using the Benjamini-Hochberg procedure [6]. This may be because of the heterogeneity of lesion locations and sizes across both groups. In contrast, the F-test is sensitive to significant differences in variance between the two groups and does show regions where the Jacobian determinant is significantly different, even after FDR correction (*q* = 0.05). The voxels close to the pial surface associated with significant differences may indicate group differences in the locations of edema, hematoma, or hemorrhage and may, therefore, be associated with an an increased risk for PTE.

**Figure 2:**
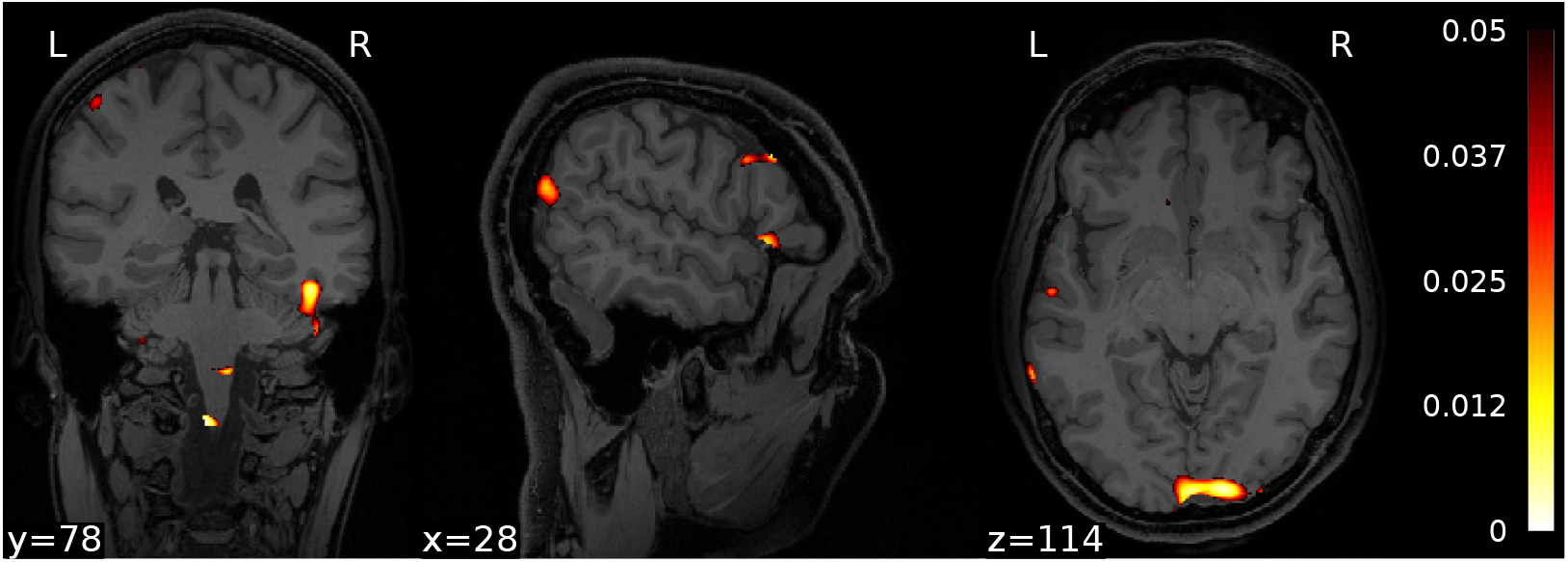
Three orthogonal views through the *t* statistic map thresholded at *p* = 0.05 (uncorrected) for TBM analysis using Jacobian determinants. No regions in the map survived multiple comparisons corrections using FDR (*q* = 0.05)

**Figure 3:**
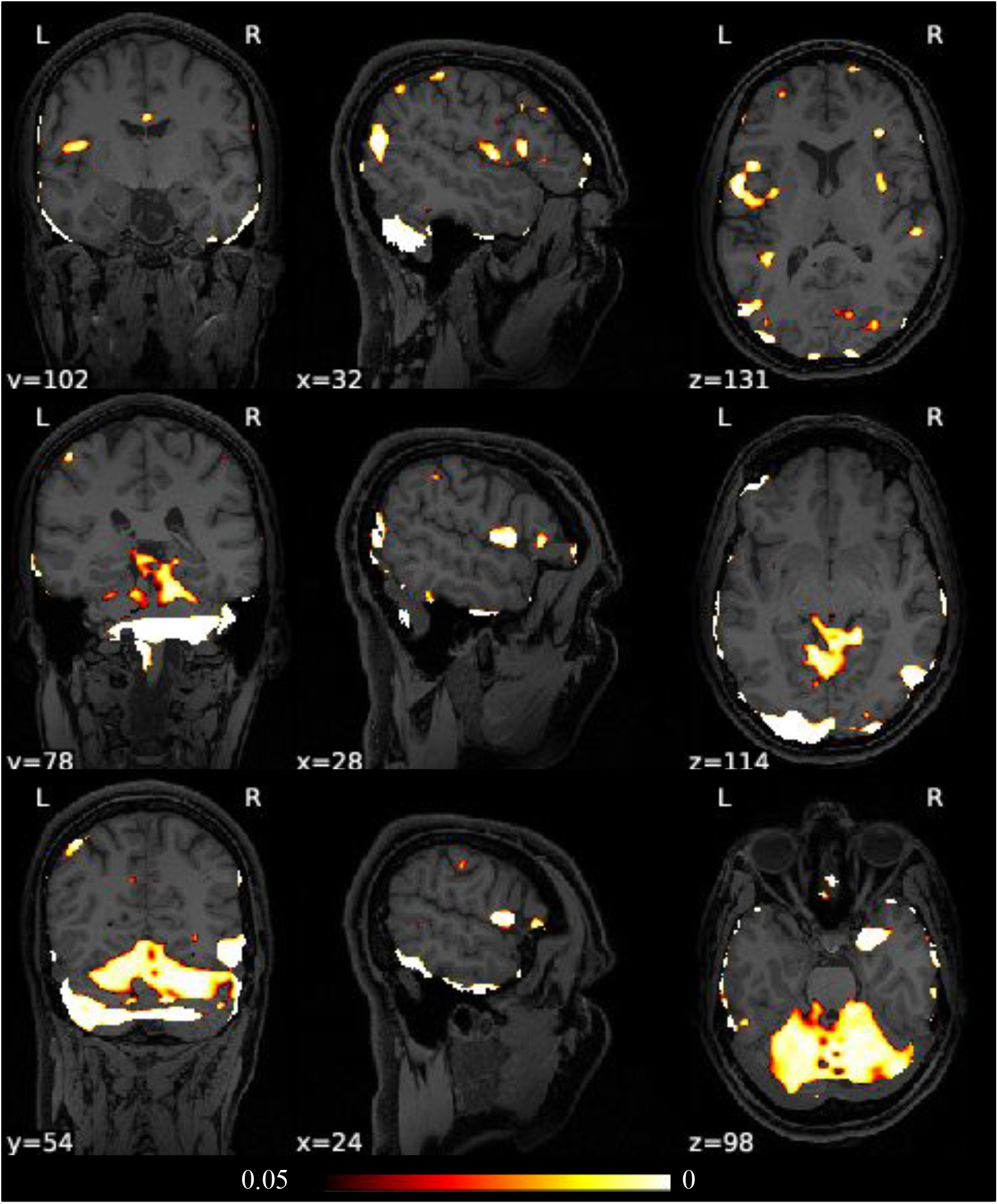
Three orthogonal views through the F-statistic map thresholded at *q* = 0.05 (FDR-corrected) for TBM analysis using Jacobian determinants.

### 3.2. Lesion-based analysis

The VAE model was trained using the independent set of 138 non-PTE training subjects, and its performance was measured using ROC analysis on the ISLEs dataset (Figure 4). Due to the infrequent occurrence of lesions in the training data, the VAE was able to prevent reconstruction of lesions so that they appeared in the error map as shown in Figure 4, where we illustrate performance for cases where lesions are present. Note that the reconstructed images in (b) are ‘de-lesioned’ approximations of the input images in (a). The normal tissues are reconstructed, whereas the anomalies and lesions are not. The error maps in (c) are indicative of anomalies in the brain. The error maps after median filtering in (d) show reasonable correspondence with the ground truth (e). The AUC for the lesion detection ROC study on the ISLEs data was 0.81. The trained model was then applied to the study population of 37 PTE and 37 non-PTE subjects.

**Figure 4:**
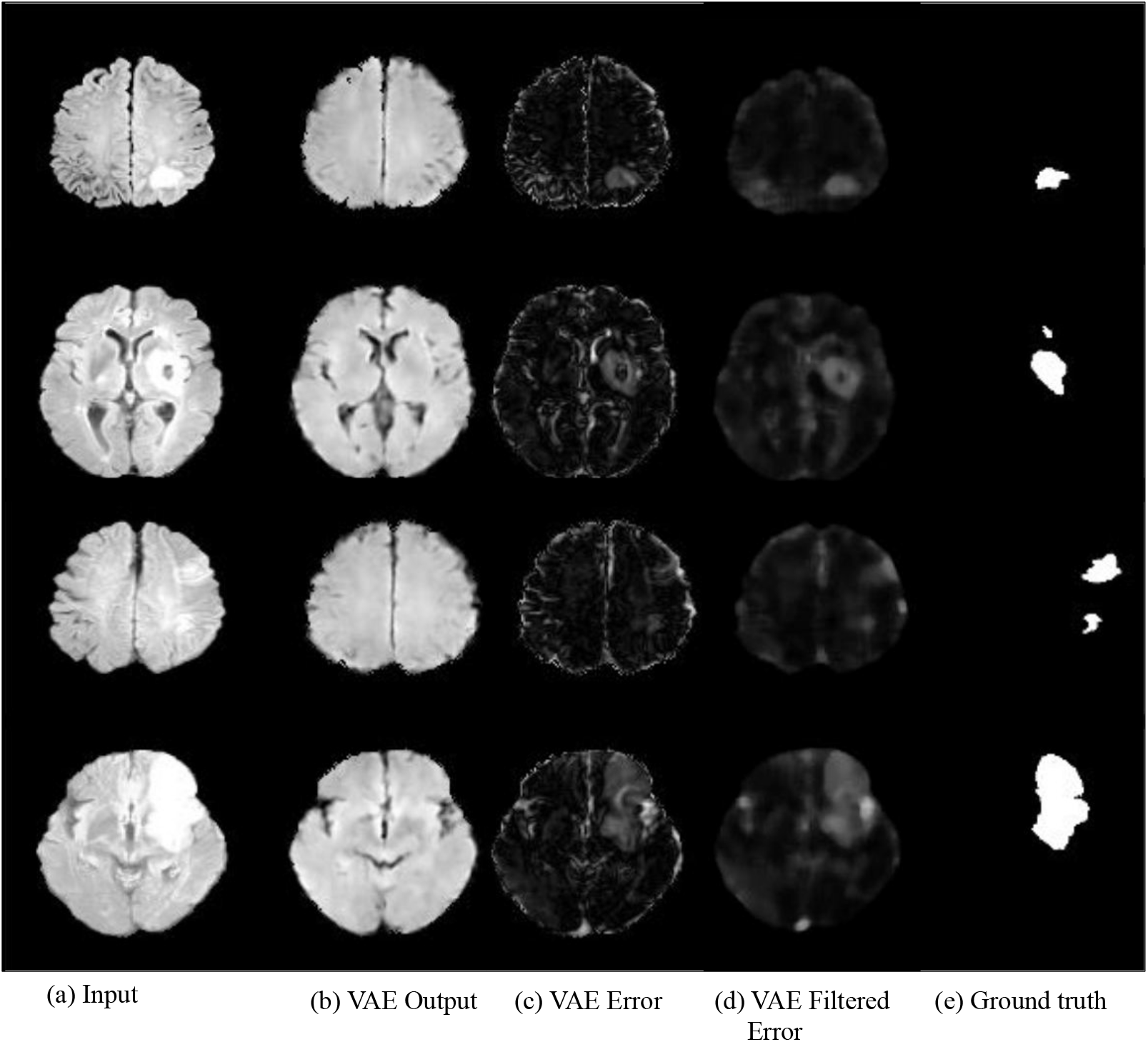
Reconstruction results obtained by applying the VAE to the ISLES dataset: (a) sample slices from input images; (b) slices reconstructed from the VAE; (c) difference between input and reconstructed images; (d) error maps after applying median filtering to reduce the occurrence of spurious voxels; (e) manually delineated lesion masks used as ground truth to evaluate VAE performance.

Mapping the results of *t*-test on VAE-computed absolute reconstruction errors shows differences in the left temporal and right frontal lobes (Figure 5). However, these results do not survive correction for multiple comparisons (*q* = 0.05). As explained earlier, this is likely because of the heterogeneity of lesions across both groups reduces the power of the *t*-test that compares the means of the two populations.

**Figure 5:**
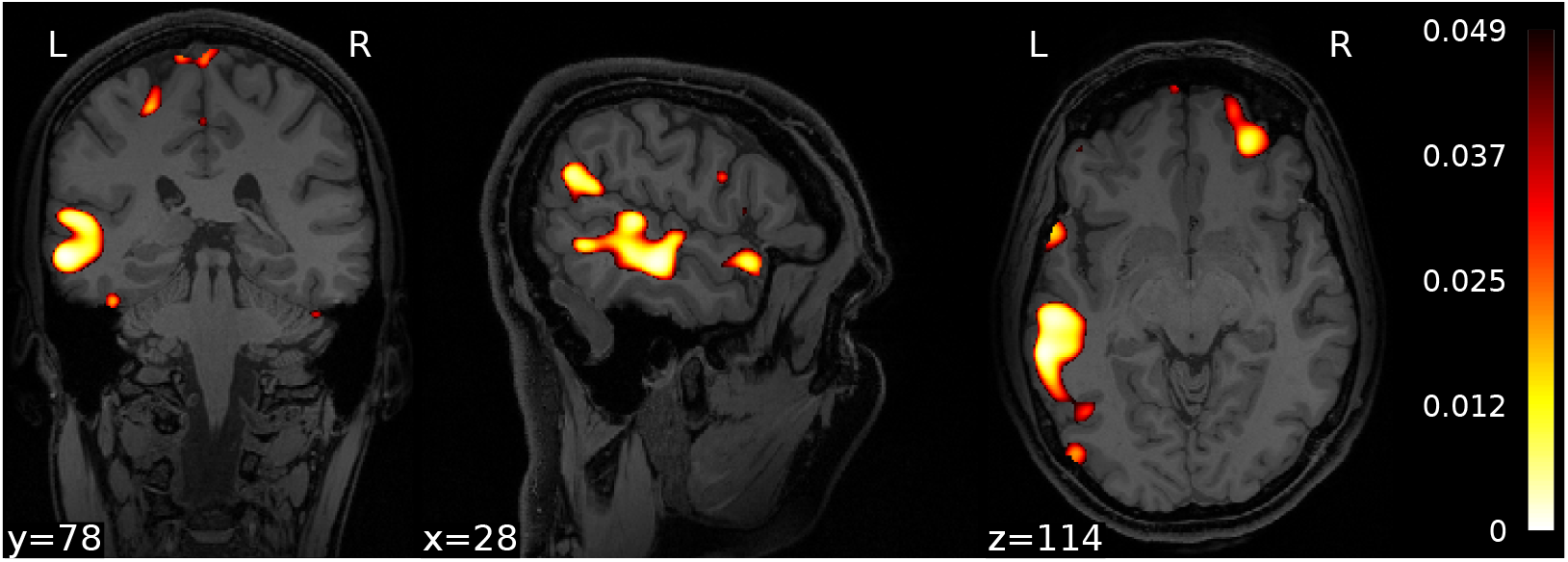
Orthogonal views through the *t*-statistic map, thresholded at *p* = 0.05 (uncorrected), comparing lesion maps for the PTE and non-PTE groups.

On the other hand, the results of the F-test showed significant differences between groups in both left and right temporal lobes. We also saw significant differences in the right occipital lobe and the cerebellum (Figure 6). The results of the lobar analysis (Table 1) were consistent with voxel-wise analysis, showing an increased variance in the PTE population relative to non-PTE subjects in the left and right temporal lobes, right occipital lobe, and cerebellum.

**Figure 6:**
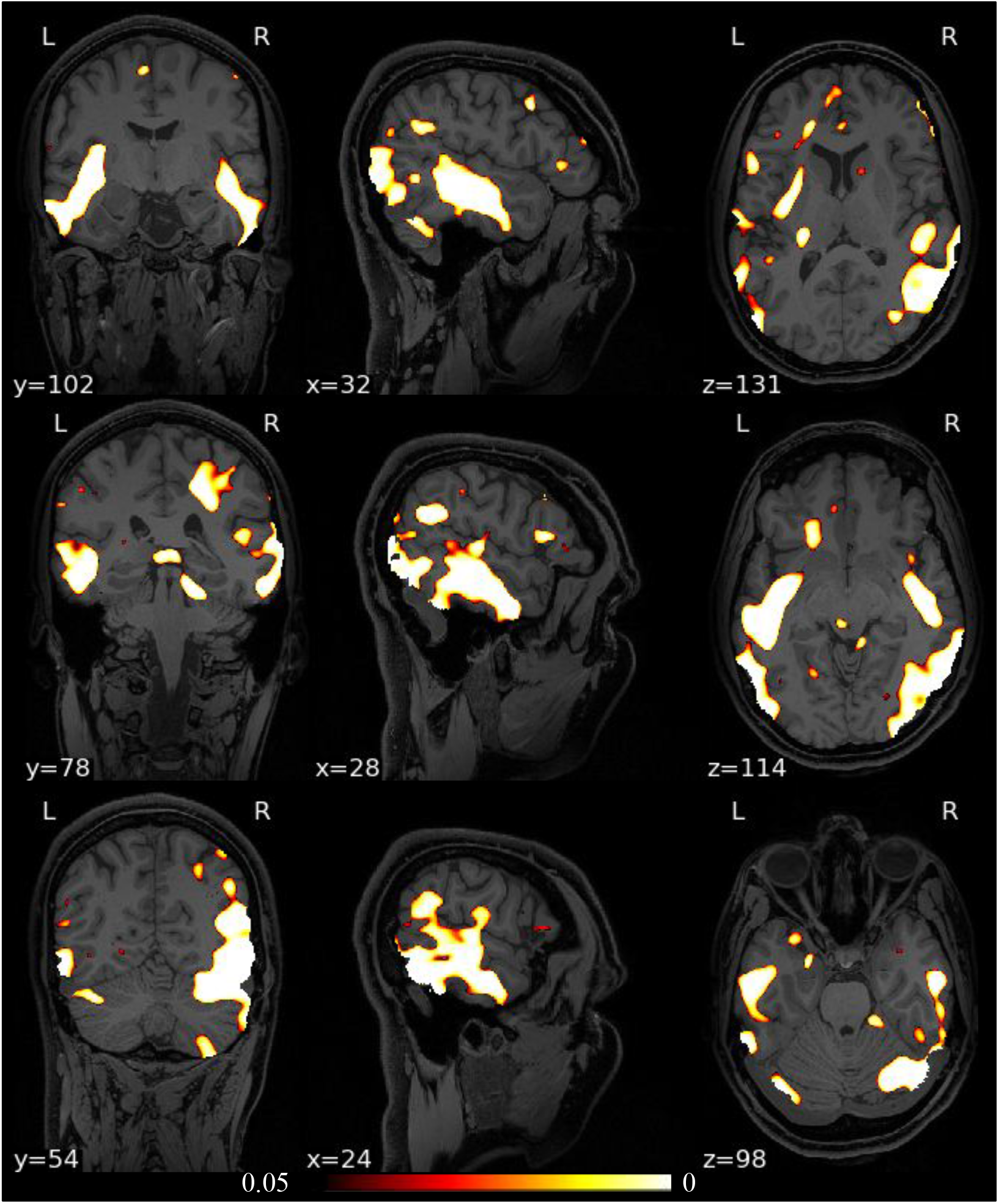
Orthogonal views through the F-statistic map, thresholded at *q* = 0.05 (FDR-corrected), comparing lesion maps for the PTE and non-PTE groups.

**Table 1:** Average lesion volumes as measured by identifying lesions using a one-class SVM on the VAE lesion maps. Red indicates cases of significant differences in the variance of lesion volume between PTE and non-PTE (F-test). The FDR-corrected *p*-values are shown at a significance level of *α* = 0.05.

| Lobe | Percentages of lobe volumes in PTE subjects median (std) | Percentages of lobe volumes in non-PTE subjects median (std) | P-value (f-test) |
| --- | --- | --- | --- |
| **Right Temporal** | 4.746 (1.496) | 4.803 (0.819) | 0.002648 |
| **Left Temporal** | 4.993 (1.223) | 4.871 (0.7661) | 0.01704 |
| **Right Occipital** | 4.131 (1.749) | 4.724 (1.225) | 0.04928 |
| **Left Occipital** | 4.336 (1.358) | 4.384 (1.001) | 0.07867 |
| **Right Frontal** | 4.794 (1.416) | 4.897 (1.868) | 0.9497 |
| **Left Frontal** | 5.251 (1.646) | 5.016 (1.399) | 0.3054 |
| **Right Parietal** | 4.794 (1.416) | 4.897 (1.868) | 0.6491 |
| **Left Parietal** | 4.610 (1.183) | 4.859 (1.320) | 0.817 |
| **Right Insula** | 5.319 (1.259) | 5.147 (1.383) | 0.6491 |
| **Left Insula** | 5.082 (1.067) | 4.612 (1.045) | 0.817 |
| **Cerebellum** | 4.758 (1.269) | 5.046 (0.8524) | 0.03532 |

## 4. Discussion and Conclusion

Our results are consistent with earlier TBI studies that show a relationship between lesion location and the probability of PTE onset. In particular, the F-test in our lesion study indicates a correlation between PTE presence and the frequency of lesion occurrence in temporal lobes, consistent with previous studies [47, 1, 48, 16, 50]. Interestingly, the TBM F-test shows areas of significant differences between groups that are, in large part, clustered on or just below the pial surface as well as in the cerebellum. Whereas the near-surface clusters could be false positives and need further investigation, this result may indicate the increased occurrence of edema or hematoma in acute TBI patients, which are known to alter cortex shape [25] and which may be associated with an increased chance for developing PTE. TBM and its extensions [5, 38] have been used for whole-brain analysis of structural abnormalities in TLE patients. Significant volume reductions were found in brain regions including the hippocampus, cingulate gyrus, precentral gyrus, right temporal lobe, and cerebellum [38]. Cross-sectional studies of children with chronic localization-related epilepsy (LRE) using traditional volumetric and voxel-based morphometry have revealed abnormalities in the cerebellum, frontal and temporal lobes, hippocampus, amygdala, and thalamus [8, 10, 18, 35, 36, 34, 49]. These studies also emphasize the role of the temporal lobe in PTE, while also providing further evidence for the role of temporal lobe lesions in PTE. The findings from our study further support this evidence for the involvement of the temporal lobe. Furthermore, studies like ours may assist or complement efforts to study post-traumatic metabolic crisis [51] or to localize post-traumatic epileptic foci for surgical resection via electroencephalography (EEG) [26, 27]. The novel use of a VAE here to automatically delineate lesions may prove useful for future studies over the large datasets, or collections of datasets like FITBIR, where manual segmentation is very time-consuming and/or subject to large inter-rater variability.

## Data Availability

The data of this meta analysis will be made available on FITBIR neuroinformatics site.

## 5. Acknowledgements

This work is supported by DoD grant W81XWH-18-1-061, and by NIH grants R01 NS074980, and R01 EB026299. A.I. is supported by NIH grant R01 NS 100973, by DoD contract W81XWH-18-1-0413 and by a Hanson-Thorell Family Research Scholarship. Data used in the preparation of this article reside in the Department of Defense (DOD) and National Institutes of Health (NIH)-supported Federal Interagency Traumatic Brain Injury Research Informatics Systems (FITBIR) in FITBIR-STUDY 0000314 and FITBIR-STUDY 0000246. This manuscript reflects the views of the authors and does not reflect the opinions or views of the DoD or the NIH.

## References

[1] Adelola Adeloye and E Latunde Odeku. The radiology of missile head wounds. Clinical radiology, 22(3):312–320, 1971.

[2] Haleh Akrami, Anand Joshi, Jian Li, Sergul Aydore, and Richard Leahy. Brain lesion detection using a robust variational autoencoder and transfer learning. In Proc. ISBI, 2020, 2020.

[3] Haleh Akrami, Anand A Joshi, Jian Li, and Richard M Leahy. Robust variational autoencoder. arXiv preprint 1905.09961, 2019.

[4] F Angeleri, J Majkowski, G Cacchio, A Sobieszek, S D’acunto, R Gesuita, A Bachleda, G Polonara, L Krolicki, M Signorino, et al. Posttraumatic epilepsy risk factors: one-year prospective study after head injury. Epilepsia, 40(9):1222–1230, 1999.

[5] John Ashburner. A fast diffeomorphic image registration algorithm. NeuroImage, 38(1):95–113, October 2007.

[6] Yoav Benjamini and Daniel Yekutieli. The control of the false discovery rate in multiple testing under dependency. Annals of Statistics, 29:1165–1188, 2001.

[7] Xiaoran Chen and Ender Konukoglu. Unsupervised detection of lesions in brain mri using constrained adversarial auto-encoders. arXiv preprint 1806.04972, 2018.

[8] Francesca Cormack, David G. Gadian, Faraneh Vargha-Khadem, J. Helen Cross, Alan Connelly, and Torsten Baldeweg. Extra-hippocampal grey matter density abnormalities in paediatric mesial temporal sclerosis. NeuroImage, 27(3):635–643, September 2005.

[9] G. Curia, M. Levitt, J. S. Fender, J. W. Miller, J. Ojemann, and R. D’Ambrosio. Impact of Injury Location and Severity on Posttraumatic Epilepsy in the Rat: Role of Frontal Neocortex. Cerebral Cortex, 21(7):1574–1592, July 2011.

[10] Melita Daley, Prabha Siddarth, Jennifer Levitt, Suresh Gurbani,W. Donald Shields, Raman Sankar, Arthur Toga, and Rochelle Caplan. Amygdala volume and psychopathology in childhood complex partial seizures. Epilepsy & Behavior: E&B, 13(1):212–217, July 2008.

[11] Emily L. Dennis, Xue Hua, Julio Villalon-Reina, Lisa M. Moran, Claudia Kernan, Talin Babikian, Richard Mink, Christopher Babbitt, Jeffrey Johnson, Christopher C. Giza, Paul M. Thompson, and Robert F. Asarnow. Tensor-Based Morphometry Reveals Volumetric Deficits in Moderate=Severe Pediatric Traumatic Brain Injury. Journal of Neurotrauma, 33(9):840–852, May 2016.

[12] Ramon Diaz-Arrastia, Mark A Agostini, Christopher J Madden, and Paul C Van Ness. Posttraumatic epilepsy: the endophenotypes of a human model of epileptogenesis. Epilepsia, 50:14–20, 2009.

[13] Richard O. Duda, Peter E. Hart, and David G. Stork. Pattern classification. John Wiley & Sons, 2012.

[14] Dominique Duncan, Giuseppe Barisano, Ryan Cabeen, Farshid Sepehrband, Rachael Garner, Adebayo Braimah, Paul Vespa, Asla Pitkänen, Meng Law, and Arthur W. Toga. Analytic Tools for Post-traumatic Epileptogenesis Biomarker Search in Multimodal Dataset of an Animal Model and Human Patients. Frontiers in Neuroinformatics, 12:86, December 2018.

[15] Behzad Eftekhar, Mohammad Ali Sahraian, Banafsheh Nouralishahi, Ali Khaji, Zahra Vahabi, Mohammad Ghodsi, Hassan Araghizadeh, Mohammad Reza Soroush, Sima Karbalaei Esmaeili, and Mehdi Masoumi. Prognostic factors in the persistence of posttraumatic epilepsy afterpenetrating head injuries sustained in war. Journal of neurosurgery, 110(2):319–326, 2009.

[16] Jeffrey Englander, Tamara Bushnik, Thao T Duong, David X Cifu, Ross Zafonte, Jerry Wright, Richard Hughes, and William Bergman. Analyzing risk factors for late posttraumatic seizures: a prospective, multi-center investigation. Archives of physical medicine and rehabilitation, 84(3):365–373, 2003.

[17] Rachael Garner, Marianna La Rocca, Giuseppe Barisano, Arthur W. Toga, Dominique Duncan, and Paul Vespa. A Machine Learning Model to Predict Seizure Susceptibility from Resting-State fMRI Connectivity. In 2019 Spring Simulation Conference (SpringSim), pages 1–11, Tucson, AZ, USA, April 2019. IEEE.

[18] Catarina A. Guimarães, Leonardo Bonilha, Renata C. Franzon, Li M. Li, Fernando Cendes, and Marilisa M. Guerreiro. Distribution of regional gray matter abnormalities in a pediatric population with temporal lobe epilepsy and correlation with neuropsychological performance. Epilepsy & Behavior: E&B, 11(4):558–566, December 2007.

[19] Rao P Gullapalli. Investigation of prognostic ability of novel imaging markers for traumatic brain injury (tbi). Technical report, BALTIMORE UNIV MD, 2011.

[20] Susan T. Herman. Epilepsy after brain insult: targeting epileptogenesis. Neurology, 59(9 Suppl 5):S21–26, November 2002.

[21] Xue Hua, Suh Lee, Igor Yanovsky, Alex D. Leow, Yi Yu Chou, April J. Ho, Boris Gutman, Arthur W. Toga, Clifford R. Jack, Matt A. Bernstein, Eric M. Reiman, Danielle J. Harvey, John Kornak, Norbert Schuff, Gene E. Alexander, Michael W. Weiner, and Paul M. Thompson. Optimizing power to track brain degeneration in Alzheimer’s disease and mild cognitive impairment with tensor-based morphometry: An ADNI study of 515 subjects. NeuroImage, 48(4):668–681, 2009.

[22] Xue Hua, Alex D Leow, Neelroop Parikshak, Suh Lee, Ming-Chang Chiang, Arthur W Toga, Clifford R Jack Jr, Michael W Weiner, Paul M Thompson, Alzheimer’s Disease Neuroimaging Initiative, et al. Tensor-based morphometry as a neuroimaging biomarker for alzheimer’s disease: an mri study of 676 ad, mci, and normal subjects. Neuroimage, 43(3):458–469, 2008.

[23] Riikka Immonen, Irina Kharatishvili, Olli Gröhn, and Asla Pitkänen. MRI Biomarkers for Post-Traumatic Epileptogenesis. Journal of Neurotrauma, 30(14):1305–1309, July 2013.

[24] Irimia. Neuroimaging of structural pathology and connectomics in traumatic brain injury: Toward personalized outcome prediction. NeuroImage: Clinical, 1:1–17” 2012.

[25] Irimia, S.Y. Goh, C.M. Torgerson, P. Vespa, and J.D. Van Horn. Structural and connectomic neuroimaging for the personalized study of longitudinal alterations in cortical shape, thickness and connectivity after traumatic brain injury. J Neurosurg Sci, 58:129–144, 2014.

[26] Andrei Irimia, SY Matthew Goh, Micah C Chambers, Carinna M Torgerson, Nathan R Stein, Paul M Vespa, and John D Van Horn. Electroencephalographic inverse localization of brain activity in acute traumatic brain injury as a guide to surgery, monitoring and treatment. Clinical Neurology and Neurosurgery, 115:2159–2165, 2013.

[27] Andrei Irimia and John D Van Horn. Epileptogenic focus localization in pharmacologically resistant post-traumatic epilepsy. Journal of Clinical Neuroscience, 11:627–631, 2015.

[28] Anand A. Joshi, David W. Shattuck, Paul M. Thompson, and Richard M. Leahy. A framework for registration, statistical characterization and classification of cortically constrained functional imaging data. In Inf Process Med Imaging, volume 19, pages 186–196, July 2005.

[29] Anand A Joshi, David W Shattuck, Paul M Thompson, and Richard M Leahy. Surface-Constrained Volumetric Brain Registration Using Harmonic Mappings. IEEE Trans. Med. Imaging, 26(12):1657–1669, 2007.

[30] Konstantinos Kamnitsas, Christian Ledig, Virginia FJ Newcombe, Joanna P Simpson, Andrew D Kane, David K Menon, Daniel Rueckert, and Ben Glocker. Efficient multi-scale 3d cnn with fully connected crf for accurate brain lesion segmentation. Medical image analysis, 36:61–78, 2017.

[31] Diederik P Kingma and Max Welling. Autoencoding variational Bayes. arXiv preprint 1312.6114, 2013.

[32] Anders Boesen Lindbo Larsen, Søren Kaae Sønderby, Hugo Larochelle, and Ole Winther. Autoencoding beyond pixels using a learned similarity metric. arXiv preprint 1512.09300, 2015.

[33] Daniel Laskowitz and Gerald Grant. Translational research in traumatic brain injury, volume 57. CRC Press, 2016.

[34] J. A. Lawson, M. J. Cook, A. F. Bleasel, V. Nayanar, K. F. Morris, and A. M. Bye. Quantitative MRI in outpatient childhood epilepsy. Epilepsia, 38(12):1289–1293, December 1997.

[35] J. A. Lawson, W. Nguyen, A. F. Bleasel, J. K. Pereira, S. Vogrin, M. J. Cook, and A. M. Bye. ILAE-defined epilepsy syndromes in children: correlation with quantitative MRI. Epilepsia, 39(12):1345–1349, December 1998.

[36] J. A. Lawson, S. Vogrin, A. F. Bleasel, M. J. Cook, L. Burns,L. McAnally, J. Pereira, and A. M. Bye. Predictors of hippocampal, cerebral, and cerebellar volume reduction in childhood epilepsy. Epilepsia, 41(12):1540–1545, December 2000.

[37] Hongwei Li, Gongfa Jiang, Jianguo Zhang, Ruixuan Wang, Zhaolei Wang, Wei-Shi Zheng, and Bjoern Menze. Fully convolutional network ensembles for white matter hyperintensities segmentation in mr images. NeuroImage, 183:650–665, 2018.

[38] Wenjing Li, Huiguang He, Jingjing Lu, Bin Lv, Meng Li, and Zhengyu Jin. Detection of whole-brain abnormalities in temporal lobe epilepsy using tensor-based morphometry with dartel. In MIPPR 2009: MedicalImaging, Parallel Processing of Images, and Optimization Techniques, volume 7497, page 749723. International Society for Optics and Photonics, 2009.

[39] Oskar Maier, Bjoern H Menze, Janina von der Gablentz, Levin Häni, Mattias P Heinrich, Matthias Liebrand, Stefan Winzeck, Abdul Basit, Paul Bentley, Liang Chen, et al. ISLES 2015-a public evaluation benchmark for ischemic stroke lesion segmentation from multispectral MRI. Medical image analysis, 35:250–269, 2017.

[40] W. Penfield. Symposium on posttraumatic epilepsy: introduction. Epilepsia, 2:109–110, 1961.

[41] Sérgio Pereira, Adriano Pinto, Victor Alves, and Carlos A Silva. Brain tumor segmentation using convolutional neural networks in mri images. IEEE transactions on medical imaging, 35(5):1240–1251, 2016.

[42] Asla Pitkänen, Tamuna Bolkvadze, and Riikka Immonen. Antiepileptogenesis in rodent post-traumatic epilepsy models. Neuroscience letters, 497(3):163–171, 2011.

[43] Asla Pitkänen and Tamuna Bolkvadze. Head Trauma and Epilepsy. In Jeffrey L. Noebels, Massimo Avoli, Michael A. Rogawski, Richard W. Olsen, and Antonio V. Delgado-Escueta, editors, Jasper’s Basic Mechanisms of the Epilepsies. National Center for Biotechnology Information (US), Bethesda (MD), 4th edition, 2012.

[44] Asla Pitkänen, Wolfgang Löscher, Annamaria Vezzani, Albert J Becker, Michele Simonato, Katarzyna Lukasiuk, Olli Gröhn, Jens P Bankstahl,Alon Friedman, Eleonora Aronica, Jan A Gorter, Teresa Ravizza, Sanjay M Sisodiya, Merab Kokaia, and Heinz Beck. Advances in the development of biomarkers for epilepsy. The Lancet Neurology, 15(8):843–856, July 2016.

[45] Dorian Pustina, Brian Avants, Michael Sperling, Richard Gorniak, Xiaosong He, Gaelle Doucet, Paul Barnett, Scott Mintzer, Ashwini Sharan, and Joseph Tracy. Predicting the laterality of temporal lobe epilepsy from PET, MRI, and DTI: A multimodal study. NeuroImage: Clinical, 9:20–31, 2015.

[46] V. Raymont, A. M. Salazar, R. Lipsky, D. Goldman, G. Tasick, and J. Grafman. Correlates of posttraumatic epilepsy 35 years following combat brain injury. Neurology, 75(3):224–229, July 2010.

[47] V Raymont, Andres M Salazar, R Lipsky, David Goldman, G Tasick, and Jordan Grafman. Correlates of posttraumatic epilepsy 35 years following combat brain injury. Neurology, 75(3):224–229, 2010.

[48] W Ritchie Russell. Disability caused by brain wounds: a review of 1,166 cases. Journal of neurology, neurosurgery, and psychiatry, 14(1):35, 1951.

[49] Duygu Tosun, Kevin Dabbs, Rochelle Caplan, Prabha Siddarth, Arthur Toga, Michael Seidenberg, and Bruce Hermann. Deformation-based morphometry of prospective neurodevelopmental changes in new onset paediatric epilepsy. Brain, 134(4):1003–1014, April 2011.

[50] Meral A Tubi, Evan Lutkenhoff, Manuel Buitrago Blanco, David McArthur, Pablo Villablanca, Benjamin Ellingson, Ramon Diaz-Arrastia, Paul Van Ness, Courtney Real, Vikesh Shrestha, et al. Early seizures and temporal lobe trauma predict post-traumatic epilepsy: A longitudinal study. Neurobiology of disease, 123:115–121, 2019.

[51] Matthew J Wright, David R McArthur, Jeffry R Alger, John D Van Horn, Andrei Irimia, Maria Filippou, Thomas C Glenn, David A Hovda, and Paul M Vespa. Early metabolic crisis-related brain atrophy and cognition in traumatic brain injury. Brain Imaging and Behavior, 7:307–315, 2013.

[52] John K Yue, Mary J Vassar, Hester F Lingsma, Shelly R Cooper, David O Okonkwo, Alex B Valadka, Wayne A Gordon, Andrew IR Maas, Pratik Mukherjee, Esther L Yuh, et al. Transforming research and clinical knowledge in traumatic brain injury pilot: multicenter implementation of the common data elements for traumatic brain injury. Journal of neurotrauma, 30(22):1831–1844, 2013.

[53] Jianguo Zhang, Kai-Kuang Ma, Meng-Hwa Er, and Vincent Chong. Tumor segmentation from magnetic resonance imaging by learning via one-class support vector machine. 2004.

